# Clinical, genomic, and neurophysiological correlates of lifetime suicide attempts among individuals with an alcohol use disorder

**DOI:** 10.1101/2023.04.28.23289173

**Authors:** Peter B. Barr, Zoe Neale, Chris Chatzinakos, Jessica Schulman, Niamh Mullins, Jian Zhang, David B. Chorlian, Chella Kamarajan, Sivan Kinreich, Ashwini K. Pandey, Gayathri Pandey, Stacey Saenz de Viteri, Laura Acion, Lance Bauer, Kathleen K. Bucholz, Grace Chan, Danielle M. Dick, Howard J. Edenberg, Tatiana Foroud, Alison Goate, Victor Hesselbrock, Emma C. Johnson, John Kramer, Dongbing Lai, Martin H. Plawecki, Jessica E. Salvatore, Leah Wetherill, Arpana Agrawal, Bernice Porjesz, Jacquelyn L. Meyers

## Abstract

Research has identified clinical, genomic, and neurophysiological markers associated with suicide attempts (SA) among individuals with psychiatric illness. However, there is limited research among those with an alcohol use disorder (AUD), despite their disproportionately higher rates of SA. We examined lifetime SA in 4,068 individuals with DSM-IV alcohol dependence from the Collaborative Study on the Genetics of Alcoholism (23% lifetime suicide attempt; 53% female; mean age: 38). Within participants with an AUD diagnosis, we explored risk across other clinical conditions, polygenic scores (PGS) for comorbid psychiatric problems, and neurocognitive functioning for lifetime suicide attempt. Participants with an AUD who had attempted suicide had greater rates of trauma exposure, major depressive disorder, post-traumatic stress disorder, and other substance use disorders compared to those who had not attempted suicide. Polygenic scores for suicide attempt, depression, and PTSD were associated with reporting a suicide attempt (ORs = 1.22 – 1.44). Participants who reported a SA also had decreased right hemispheric frontal-parietal theta and decreased interhemispheric temporal-parietal alpha electroencephalogram resting-state coherences relative to those who did not, but differences were small. Overall, individuals with an AUD who report a lifetime suicide attempt appear to experience greater levels of trauma, have more severe comorbidities, and carry polygenic risk for a variety of psychiatric problems. Our results demonstrate the need to further investigate suicide attempts in the presence of substance use disorders.

## Introduction

Approximately 2-5% of U.S. adults report having attempted suicide in their lifetimes (Baca-Garcia et al., 2010; Kessler et al., 1999; Scheer et al., 2020), with the prevalence increasing in more recent birth cohorts (Olfson et al., 2017). Additionally, deaths by suicide are one of the leading causes in the recent decline in U.S. life expectancy, alongside other “deaths of despair” such as drug and alcohol related deaths (Case & Deaton, 2015; Tilstra et al., 2021). While the rate of suicide attempts in the general population is alarming, the rate of lifetime suicide attempts is greater than triple (17.5%) among those with an alcohol use disorder (AUD) (Potash et al., 2000). Among those seeking treatment for AUD, 40% report at least one suicide attempt at some point in their lives (Koller et al., 2002; Modesto-Lowe et al., 2006; Sher, 2006; Whiteford et al., 2013). A history of past suicide attempts is among the most prominent predictors of subsequent suicide death and contributes significant health care and disability costs per attempt (Shepard et al., 2016). Research focused on correlates of suicide attempts can potentially help identify and treat those with non-fatal suicide attempts, with the goal of reducing suicide deaths and saving lives (Yuodelis-Flores & Ries, 2015). Importantly, alcohol use is a consistent risk factor for death by suicide (Isaacs et al., 2022), individuals with an AUD have emerged as a particularly high-risk group (Edwards et al., 2024; Lannoy et al., 2022, 2024).

In addition to clinical and phenotypic correlations between substance use disorders (SUD), and suicidal behaviors there is also consistent evidence for genetic overlap between these outcomes, with AUD in particular. Evidence from large scale genome-wide association studies (GWAS) of both AUD/problematic alcohol use (PAU) (Kranzler et al., 2019; Sanchez-Roige et al., 2019; Walters et al., 2018; Zhou et al., 2023) and suicide attempts (SA) (Docherty et al., 2023; Mullins et al., 2022), reveal robust genetic correlations across these outcomes. And while there are very limited GWASs of suicide attempt in the presence of AUD (Peng et al., 2024), two recent efforts using multivariate approaches which harness existing GWAS have shown that: 1) a shared genetic liability towards all forms of SUD is correlated with suicidal ideation, attempt, and self-injurious behavior, independent of genetic liability towards depression (Colbert et al., 2021), and 2) the shared genetic overlap between AUD and suicide attempts is explained, in part, by underlying liability towards impulsive behaviors (Stephenson et al., 2023).

Similar to the genetics of AUD and SA, two separate literatures have explored neurocognitive differences between (a) individuals who have attempted suicide to those who have not (Keilp et al., 2013; Richard-Devantoy et al., 2012, 2014) and (b) individuals with AUD (Cabé et al., 2016; Le Berre, 2019; Le Berre et al., 2017) compared to those unaffected with AUD. Those with AUD exhibit deficits in many domains of brain functioning, including neuropsychological performance, and neurophysiological indices (Cabé et al., 2016; Le Berre, 2019; Le Berre et al., 2017). Executive functioning is typically the primary focus of such studies, with a large literature demonstrating that individuals with AUD display poorer executive functioning and atypical neurophysiological profiles (e.g., EEG connectivity) than individuals without AUD (Cardenas et al., 2018; Kamarajan et al., 2020; Mumtaz et al., 2017; Park et al., 2017). Researchers have also examined these areas of brain functioning among individuals who have exhibited suicidal ideation and related mental health problems, such as depression (Keilp et al., 2013; Richard-Devantoy et al., 2012, 2014), though research focused on SA is limited. While no previous studies have examined EEG connectivity and SA, EEG connectivity in depressed patients exhibited higher alpha and theta coherences in frontal, temporal, and parietal regions, and higher beta coherence in frontal and temporal regions (Leuchter et al., 2012a). Further, a recent study found other neurophysiological differences associated with binge drinking and suicidal behaviors in adolescents (Ehlers et al., 2020). To our knowledge, no prior study has examined neural connectivity among those with AUD who have attempted suicide.

Given the higher rates of SA observed among those with AUD, we explored whether there are clinical, genomic, and neurophysiological markers of SA within this population. Among participants diagnosed with an AUD (DSM-IV alcohol dependence) drawn from the Collaborative Study on the Genetics of Alcoholism (COGA), we examined whether clinical risk factors, polygenic scores (PGS) for comorbid psychiatric problems, and neurocognitive functioning differed between those who have and have not reported a lifetime suicide attempt.

## Methods

### Sample and Measures

The Collaborative Study on the Genetics of Alcoholism (COGA) is a large, multi-site study of 2,255 families affected with AUD, designed to identify and understand genetic factors involved in the predisposition to AUD and related disorders, as previously described (Agrawal et al., 2023; Begleiter, 1995; Dick et al., 2023). Probands along with all willing first-degree relatives were assessed; recruitment was extended to include additional relatives in families that contained 2 or more first degree relatives with alcohol dependence and community ascertained comparison families (N = 17,878). Participants 18 or older completed the Semi-Structured Assessment for the Genetics of Alcoholism (SSAGA) which is a poly-diagnostic interview (Bucholz et al., 1994), and participants ages 12-17 completed an adolescent SSAGA. We currently have genome wide data on 12,145 individuals. Our final analytic sample consisted of 4,068 COGA participants with an alcohol dependence diagnosis (lifetime) and GWAS data (including 3,270 individuals of European-like and 798 individuals of African-like genetic similarity, see following section for discussion on assignment of genetic similarity).

### Lifetime suicide attempt

All participants were queried about whether they had “ever tried to kill” themselves (*suicide attempt*), regardless of a history of suicidal ideation (i.e., thoughts about killing yourself). For the current analyses, we included individuals reporting any suicide attempt, including those reporting drug-related suicide attempt (14% of all attempts). Importantly, suicide attempt items were not exclusively nested within the diagnostic section for major depressive disorder (MDD), although individuals who reported suicide attempts in that section were coded accordingly as having reported the behavior.

### Clinical Risk Factors and Comorbidities

We created lifetime diagnoses of other substance use disorders (SUD), psychiatric disorders, suicidal thoughts and behaviors, and trauma exposure based upon DSM-IV criteria using the child and adult versions of the SSAGA (Dick et al., 2023). We assessed nicotine dependence using the Fagerström Test for Nicotine Dependence (FTND) scores (Heatherton et al., 1991). Additionally, we included measures of extended family histories of AUD, and other alcohol-related problems (Pandey et al., 2020).

### Polygenic scores (PGS)

Genotyping, imputation and quality control have been described previously (Johnson et al., 2023; Lai et al., 2021). Briefly, in order to limit the impact of population structure, genetic data were used to assign individuals into genetically similar groupings (National Academies of Sciences and Medicine, 2023) based on the first two principal components and the 1000 genomes reference panel (Phase 3, version 5) (Johnson et al., 2023). Families were classified as primarily European-like (EUR-like) or African-like (AFR-like) according to the genetic similarity of the greatest proportion of family members (Lai et al., 2021). Genotyping of individuals in the analytic sample was performed using the Illumina 2.5M array (Illumina, San Diego, CA, USA), the Illumina OmniExpress (Wang et al., 2013), or the Illumina 1M array, or the Affymetrix Smokescreen array (Baurley et al., 2016). SNPs with a genotyping rate <98%, Hardy-Weinberg equilibrium violations (p<10^-6^), or with minor allele frequency (MAF) less than 3% were excluded from analyses. Data were imputed to 1000 genomes (Phase 3) using SHAPEIT (Delaneau et al., 2013) and IMPUTE2 (Das et al., 2016). Following imputation, dosage probabilities ≥ 0.90 were converted to hard calls. SNPs with an imputation information score < 0.30 or MAF < 0.03 were excluded from subsequent analysis.

We estimated polygenic scores (PGS), which are aggregate measures of the number of risk alleles individuals carry weighted by effect sizes from GWAS summary statistics, for a variety of psychiatric and substance use phenotypes. We included PGS derived from recent GWAS of (1) alcohol use disorders (AUD) (Zhou et al., 2020), (2) depression (DEP, 23andMe excluded) (Levey et al., 2021), (3) post-traumatic stress disorder (PTSD) (Nievergelt et al., 2019), (4) bipolar disorders (BIP) (Bigdeli et al., 2020; Mullins et al., 2021), (5) schizophrenia (SCZ) (Bigdeli et al., 2020; Trubetskoy et al., 2022) (6) smoking initiation (SMOK, as a proxy for externalizing risk) (Karlsson Linnér et al., 2021; Saunders et al., 2022) and (7) suicide attempt (SUI) (Docherty et al., 2023). For AUD and BIP, we meta-analyzed published GWAS results with corresponding results from FinnGen (release 9) (Kurki et al., 2023). We focus on these PGS specifically because: 1) these disorders are phenotypically correlated with suicide attempt, and 2) they contain GWAS results for both European-like and African-like groupings. For GWAS that originally included COGA in the discovery sample, we obtained summary statistics with COGA removed.

To date, GWAS have been overwhelmingly limited to individuals of primarily European descent (Mills & Rahal, 2019). Because of variation in allele frequencies and linkage disequilibrium (LD) patterns, PGS often lose predictive accuracy when there is mismatch between the genetic similarity of the discovery GWAS and target sample (Ding et al., 2023). COGA includes participants of both African-like and European-like groupings, thus we used PRS-CSx (Ruan et al., 2022), a method that integrates GWAS summary statistics from well-powered GWAS (typically of European-like individuals) with those from other populations to improve the predictive power of PGS in the participants of African-like groupings in COGA. PRS-CSx employs a Bayesian approach to correct GWAS summary statistics for the non-independence of SNPs in LD. We converted PGS into Z-scores for ease of interpretation

### Electroencephalogram (EEG) data

EEG recording and processing have been detailed previously (Meyers et al., 2023). Briefly, resting (eyes-closed) EEG was recorded for 4.25 min; a continuous interval of 256 seconds was analyzed. Each subject wore a fitted electrode cap using the 61-channel montage as specified according to the extended 10–20 International system. The nose served as reference and the ground electrode was placed on the forehead. Electrode impedances were always maintained below 5 kΩ. EEG was recorded with subjects seated comfortably in a dimly lit sound-attenuated temperature-regulated booth. They were instructed to keep their eyes closed and remain relaxed, but not to fall asleep. Electrical activity was amplified 10,000 times by Neuroscan and Masscomp amplifiers, with a bandpass between 0.02 Hz to 100 Hz and recorded using the Neuroscan system (Compumedics Limited; El Paso, TX). EEG procedures were identical at all COGA collection sites. Bipolar electrode pairs were derived to reduce volume conduction effects, and 27 representative coherence pairs were selected based on previous EEG coherence work in COGA (Meyers et al., 2023). Magnitude squared coherence was calculated from power spectral values derived from Fourier Conventional Fourier transform methods (Nunez et al., 1997). Coherence measures were generated between bipolar pairs at the following frequency bands: theta (3-7 Hz), alpha (7-12 Hz), beta (12-28 Hz).

### Statistical analyses

We compared those with an AUD who reported a suicide attempt and those with an AUD who did not report a suicide attempt across a range of sociodemographic, psychiatric comorbidities, experiences of trauma, and other measures related to alcohol misuse. We use multiple-group, multi-level regression models in Mplus (Rattana Wannisa & Rattana Narunart, 2558) and adjusted for sex, age (at time of psychiatric assessment), genetic similarity (AFR-like vs. EUR-like), family history of AUD, and family relatedness. We ran all models simultaneously (i.e., correlation among all variables accounted for) to limit multiple testing.

For polygenic scores, we first compared those with AUD who had reported a suicide attempt to those with AUD who had not reported a suicide attempt across all PGSs, independently, using logistic regression in R (version 4.2.1). Second, to ensure that results within those with AUD were not biased by conditioning on AUD (Akimova et al., 2021), we also compared: 1) those with AUD who had a reported suicide attempt, 2) those with AUD who had not reported a suicide attempt, and 3) those without AUD who had a reported suicide attempt to those who neither reported a suicide attempt nor meet criteria for AUD (see Supplemental Table 1 for sample description) using a multinomial logistic regression model in the *nnet* package in R (Ripley & Venables, 2022). In both analyses, we included sex, age, the first six genetic principal components (PCs), genotype array, and birth cohort as covariates. To adjust for familial clustering, we used cluster robust standard errors (Colin Cameron et al., 2011; Davenport et al., 2011). We stratified analyses by genetic similarity and then meta-analyzed results (by PGS) within each of the analyses above. All analyses were corrected for multiple testing. We also performed a GWAS of SA, but lacked the power to identify any individual variants associated with SA (see supplementary information)

Lastly, we compared those with AUD who reported a suicide attempt and those with AUD who did not report a suicide attempt across neurophysiological measures (resting state EEG coherence), again using multiple-group, multi-level regression models adjusted for sex, age (at time of psychiatric assessment), genetic similarity, family history of AUD, and family relatedness. We ran all models simultaneously to limit multiple testing.). We also performed a series of exploratory analyses within a subset of individuals who had available neurocognitive measures (see supplemental information for full description).

## Results

### Clinical Risk Factors Associated with Suicide Attempt in Participants with AUD

The main analytic sample was limited to the 4,068 participants with a DSM-IV diagnoses of alcohol dependence. We compared 3,138 COGA participants who met criteria for AUD and did not attempt suicide in their lifetime with 930 participants with AUD who attempted suicide. Table 1 presents the demographic characteristics. Overall, those with AUD who attempted suicide were more likely to be female (53% vs. 32%). Rates of suicide attempt and the age distribution of participants were similar across EUR-like and AFR-like groups (see Table S2 for stratified results). The majority (58.4%) of the analytic sample endorsed suicidal ideation at some point in their lifetime; of those who attempted suicide, 97.6% endorsed prior suicidal ideation compared to 46.8% of those who did not attempt suicide.

**Table 1.** Sociodemographic Characteristics in COGA Participants with an Alcohol Use Disorder (N = 4,068)

|  | No Suicide Attempt<br>(N = 3,138) | Suicide Attempt<br>(N = 930) |
| --- | --- | --- |
| Female (%) | 31.8% | 53.1%* |
| Black or African American (%) | 20.5% | 16.7% |
| Hispanic (%) | 6.2% | 8.5% |
| Mean age at last interview (SD) | 39.9 (11.9) | 38.2 (10.5) |
| Suicidal Ideation (%) | 46.8% | 97.6%* |

\* $p < .05$

Figure 1 presents the means and rates for clinical and psychiatric comorbidities cross those with and without a history of SA. Participants with AUD who had attempted suicide were significantly more likely to have been exposed to traumatic events in their life, regardless of the type of trauma (sexual, assaultive and non-assaultive). Additionally, those who reported suicide attempt also had significantly higher lifetime rates of major depressive disorder and post-traumatic stress disorder relative to those who had not attempted suicide. In terms of comorbid substance use addition, participants that reported attempting suicide had higher family history densities of AUD (Pandey et al., 2020), started drinking at an earlier age, had more severe indicators of alcohol-related problems, and had higher rates of meeting lifetime criteria for other SUDs (cocaine, nicotine, sedative, and stimulant). In total, those with AUD who report suicide attempt seem to be more severely affected for other psychiatric and substance use disorders.

**Figure 1:**
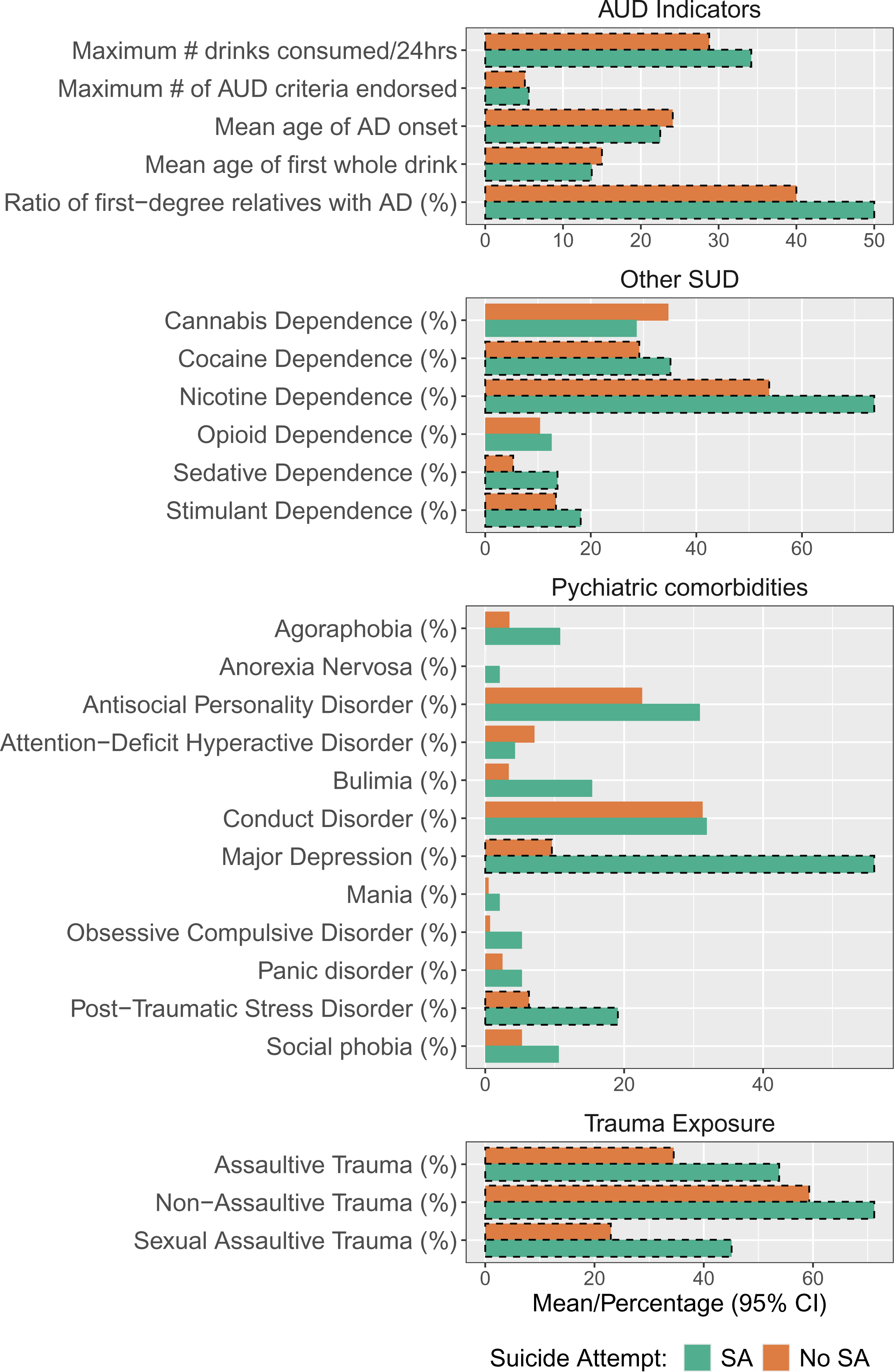
Clinical comorbidities across those who have and have not reported a suicide attempt. Percentages and means for psychiatric and substance use comorbidities for those who have attempted (SA) and have not attempted (No SA) suicide. Significant differences (*p < .*05) indicated by dashed lines around bars.

### Polygenic Scores

Figure 2, Panel A presents the meta-analyzed results for associations between each of the corresponding PGSs and lifetime suicide attempt within those meeting criteria for AUD. PGSs for DEP (OR*_META_* = 1.34, 95% CI = 1.18, 1.53), PTSD (OR*_META_* = 1.23, 95% CI = 1.03, 1.45), and SUI (OR*_META_*= 1.44, 95% CI = 1.22, 1.70) were associated with increased odds of reporting suicide attempt. However, the AUD, BIP, SCZ, and SMOK PGSs were not associated with suicide attempt (stratified results in Table S3).

**Figure 2:**
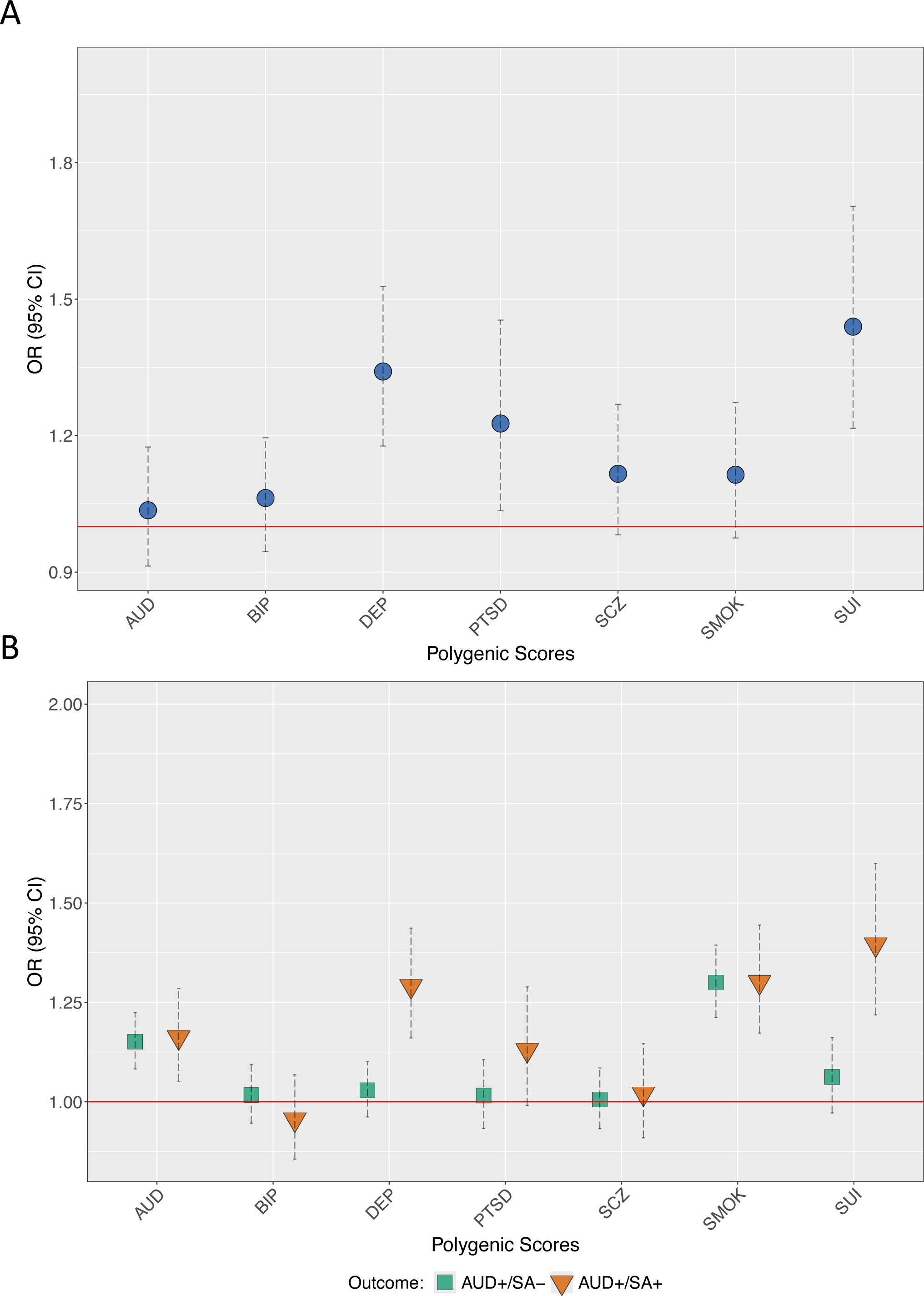
Polygenic scores across those who have and have not reported a suicide attempt. Panel A presents odds ratios (OR) for AUD, DEP, and SUI PGSs from logistic regression models in persons with AUD who had and had not attempted suicide. Panel B presents OR from multinomial logistic models (no AUD, no suicide attempt as reference group). All models include cohort, sex, PC1-PC3, array, and site as covariates. SEs adjusted for familial clustering using cluster-robust standard errors. AFR = African-like genetic similarity grouping, EUR = European like genetic similarity grouping, AUD = alcohol use disorder polygenic score, DEP = depression polygenic score, = SUI suicide attempt polygenic score, SA-= no lifetime suicide attempt, AUD-= does not meet criteria for alcohol use disorder, SA+ = lifetime suicide attempt, AUD+ = meets criteria for alcohol use disorder.

Figure 2 (Panel B) shows conditional PGS results from the multinomial logistic models comparing those with AUD who had attempted suicide (AUD+/SA+) and those with AUD who had not attempted suicide (AUD+/SA-), to those without an AUD diagnosis and who had not attempted suicide (AUD-/SA+ omitted for clarity, full results in Table S4). Relative to the AUD-/SA-group, the AUD (OR*_META_* = 1.16, 95% CI = 1.05, 1.28), DEP (OR*_META_* = 1.29, 95% CI = 1.16, 1.44), SMOK (OR*_META_* = 1.30, 95% CI = 1.17, 1.44), and SUI (OR*_META_* = 1.40, 95% CI = 1.22, 1.60) PGSs were all associated with increased odds of being in the AUD+/SA+ group. By contrast, only the AUD (OR*_META_* = 1.15, 95% CI = 1.08, 1.22) and SMOK (OR*_META_* = 1.30, 95% CI = 1.21, 1.39) PGSs were associated with increased odds of being in the AUD+/SA-group relative to the reference group (AUD-/SA-).

### Neurophysiological Findings

We observed nominal differences in resting state EEG coherence patterns in those with AUD that had attempted suicide compared to those who had not attempted suicide. However, only two findings withstood multiple test correction: decreased right hemispheric frontal-parietal theta (3-7Hz @ F8-F4--P8-P4) and decreased interhemispheric temporal-parietal alpha (7-12 Hz @ T8-P8--T7-P7) EEG resting-state coherences (p<0.001, Figure 3). Our exploratory analyses within a subset of individuals who had available neurocognitive measures did not produce any significant results (available in the supplementary information).

**Figure 3.**
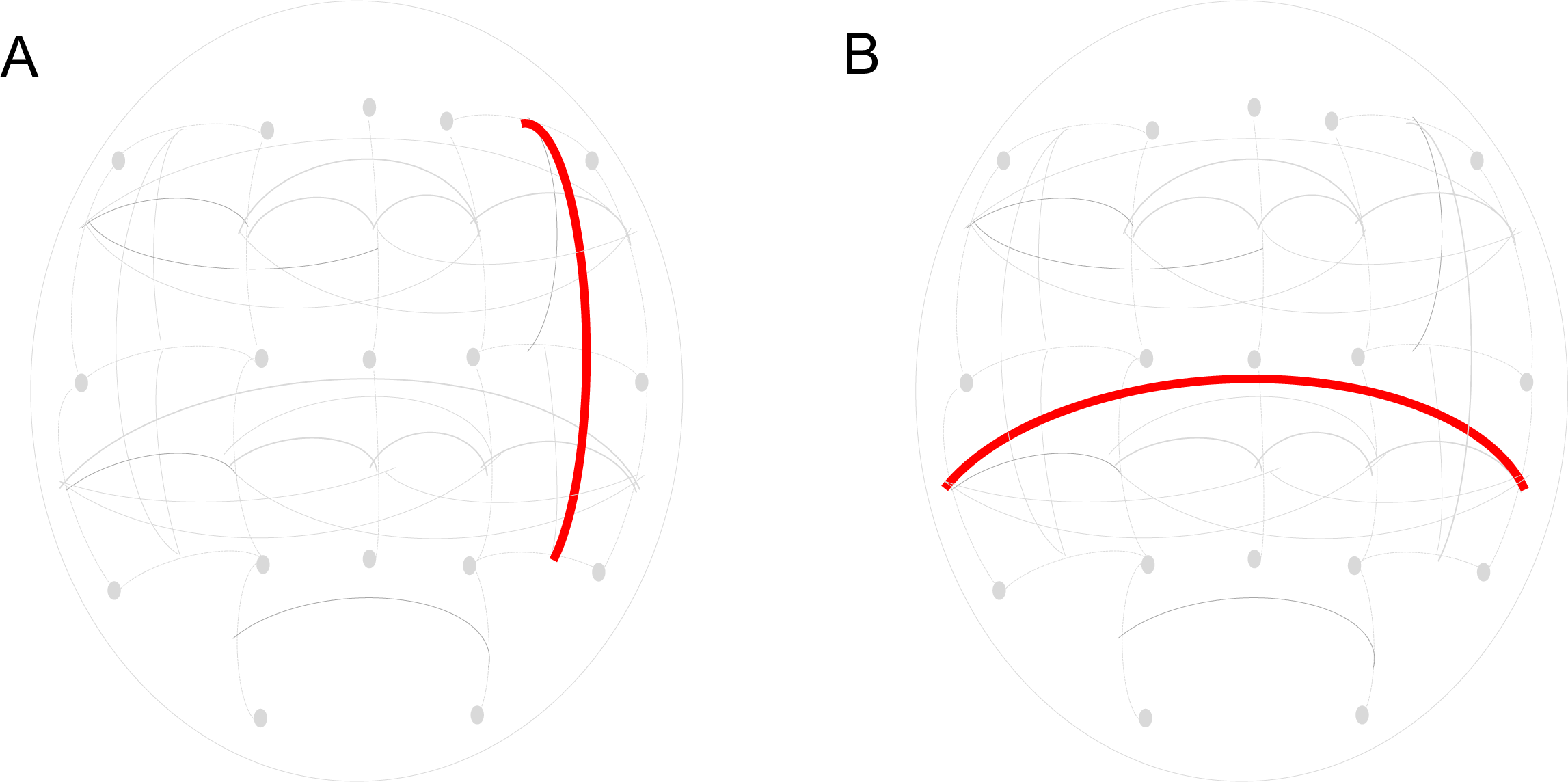
Neurophysiological measures across those with and without a reported suicide attempt. Decreased right hemispheric frontal-parietal theta (3-7 Hz @ F8-F4--P8-P4) and decreased interhemispheric temporal-parietal (7-12 Hz @ T8-P8--T7-P7) alpha EEG resting-state coherences. A) Decreased right hemispheric frontal-parietal theta (3-7 Hz @ F8-F4--P8-P4) resting-state EEG coherence. B) Decreased interhemispheric temporal-parietal alpha (7-12 Hz @ T8-P8--T7-P7) resting-state EEG coherence

## Discussion

Researchers have begun to identify clinical, genomic, and neurophysiological correlates of suicide attempts among individuals with and without psychiatric illnesses (i.e., schizophrenia, bipolar disorder, depression) [13,14,15,16]. However, few have examined these risk factors in tandem among those with AUD, despite the higher rates of suicide attempts of this group. The current study identified distinct clinical, genomic, and neurophysiological associations with lifetime suicide attempt among individuals who meet criteria for DSM-IV alcohol dependence.

All participants with an AUD in COGA reported elevated rates of suicidal ideation, other substance use disorders, and trauma exposure compared to the general population (Baca-Garcia et al., 2010; Grant et al., 2016). However, those who met criteria for an AUD and reported a lifetime suicide attempt had even greater levels of lifetime trauma (sexual, assaultive, and non-assaultive), other substance related problems, suicidal ideation, and comorbid psychiatric conditions (PTSD and major depressive disorder) relative to those who had not attempted suicide. These results confirm that those with AUD who report a lifetime suicide attempt represent clinically high-risk group. Given the strong role early trauma plays in risk for suicide attempt (Zatti et al., 2017), the elevated levels of exposure to trauma in this group, and the higher rates of PTSD, trauma exposure may play an even more important role for SA risk in those with AUD. Future work should utilize prospective information to determine whether early trauma exposure and psychiatric problems predate the onset of AUD and eventual SA in COGA.

In terms of genetic risk, the polygenic scores for suicide attempt, depression, and PTSD were associated with a lifetime suicide attempt in persons with an AUD in the meta-analyzed results. Exploration of the stratified results demonstrate these were primarily driven by the associations in the EUR-like participants. The lack of associations of PGSs in the AFR-like grouping likely stems from the relatively small sample sizes of the discovery GWASs (Dudbridge, 2013). Importantly, in the multinomial logistic regression models, those with AUD did not differ in mean levels of AUD PGS regardless of whether they had reported a lifetime suicide attempt. Similar to the logistic regression models limited to persons with AUD, those who had attempted suicide had higher suicide and depression PGSs.

We also observed significant decreases in right hemispheric frontal-parietal theta (3-7 Hz @ F8-F4--P8-P4) and interhemispheric temporal-parietal alpha (7-12 Hz @ T8-P8--T7-P7) EEG resting-state coherences in the resting state among AUD individuals who had attempted suicide. While there are no prior studies of suicide attempt that examined EEG coherence, differences in alpha and theta coherences in frontal, temporal, and parietal regions, and higher beta coherence in frontal and temporal regions were found previously in depressed patients (Leuchter et al., 2012b). Together, these data suggest that while both decreased theta and alpha resting state connectivity are likely among AUD individuals with depression and suicide attempts, but more data are needed to make definitive any conclusions.

We note several important limitations. First, we focused on lifetime risk for all of the clinical measures included and cannot speak to the time order between these clinical risk factors and suicide attempt. Future research can harness the smaller subset of prospective participants in COGA to examine the longitudinal associations between psychiatric conditions and future suicide attempt among those with an SUD. Second, we focused exclusively on AUD. There is also a high rate of suicide attempts among individuals with other SUDs, particularly cocaine and opioid use disorders. Lastly, we did not have data on those who died by suicide, which may differ from those who have attempted but not taken their own lives.

Understanding the antecedents for suicide attempt remains one of the top goals of psychiatric epidemiology. Persons with substance use disorders are a particularly at-risk group for lifetime suicide attempt. In the current analysis, we demonstrated that persons with an AUD that attempted suicide had particularly higher levels of trauma exposure and psychiatric comorbidities, elevated polygenic scores for suicide attempt, depression, and PTSD, and lower neurophysiological functioning. Future work with larger and more diverse samples can examine additional risk factors, such as social and environmental conditions. Identifying robust predictive markers within an already high risk group may allow for earlier intervention and prevention from unnecessary loss of human life.

## Data Availability

COGA data are available through the National Institute on Alcohol Abuse and Alcoholism or the database of Genotypes and Phenotypes; dbGaP(phs000763.v1.p1, phs000125.v1.p1).

## Acknowledgements

Research reported in this publication was supported by the National Institute on Alcohol Abuse and Alcoholism under award numbers U01AA008401. The content is solely the responsibility of the authors and does not necessarily represent the official views of the National Institutes of Health. This research also used summary data from the Psychiatric Genomics Consortium (PGC), the Million Veterans Program (MVP), and the International Suicide Genetics Consortium (ISGC, now part of the PGC). We would like to thank the many studies that made these consortia possible, the researchers involved, and the participants in those studies, without whom this effort would not be possible.

The Collaborative Study on the Genetics of Alcoholism (COGA), Principal Investigators B. Porjesz, V. Hesselbrock, T. Foroud; Scientific Director, A. Agrawal; Translational Director, D. Dick, includes ten different centers: University of Connecticut (V. Hesselbrock); Indiana University (H.J. Edenberg, T. Foroud, Y. Liu, M.H. Plawecki); University of Iowa Carver College of Medicine (S. Kuperman, J. Kramer); SUNY Downstate Health Sciences University (B. Porjesz, J. Meyers, C. Kamarajan, A. Pandey); Washington University in St. Louis (L. Bierut, J. Rice, K. Bucholz, A. Agrawal); University of California at San Diego (M. Schuckit); Rutgers University (J. Tischfield, D. Dick, R. Hart, J. Salvatore); The Children’s Hospital of Philadelphia, University of Pennsylvania (L. Almasy); Icahn School of Medicine at Mount Sinai (A. Goate, P. Slesinger); and Howard University (D. Scott). Other COGA collaborators include: L. Bauer (University of Connecticut); J. Nurnberger Jr., L. Wetherill, X., Xuei, D. Lai, S. O’Connor, (Indiana University); G. Chan (University of Iowa; University of Connecticut); D.B. Chorlian, J. Zhang, P. Barr, S. Kinreich, G. Pandey (SUNY Downstate); N. Mullins (Icahn School of Medicine at Mount Sinai); A. Anokhin, S. Hartz, E. Johnson, V. McCutcheon, S. Saccone (Washington University); J. Moore, F. Aliev, Z. Pang, S. Kuo (Rutgers University); A. Merikangas (The Children’s Hospital of Philadelphia and University of Pennsylvania); H. Chin and A. Parsian are the NIAAA Staff Collaborators. We continue to be inspired by our memories of Henri Begleiter and Theodore Reich, founding PI and Co-PI of COGA, and also owe a debt of gratitude to other past organizers of COGA, including Ting-Kai Li, P. Michael Conneally, Raymond Crowe, and Wendy Reich, for their critical contributions. This national collaborative study is supported by NIH Grant U10AA008401 from the National Institute on Alcohol Abuse and Alcoholism (NIAAA) and the National Institute on Drug Abuse (NIDA).

## Disclosures

The authors do not have any conflicts of interest to report.

## Data availability

All data sources are described in the manuscript and supplemental information. No new data were collected. COGA genetic data available through dbGaP (Study Accession: phs000763.v1.p1). The process for obtaining the GWAS summary statistics used in these analyses are described in the corresponding original GWAS publications.

## Notes

### Competing Interest Statement

The authors have declared no competing interest.

### Funding Statement

COGA is supported by NIH Grant U10AA008401 from the National Institute on Alcohol Abuse and Alcoholism (NIAAA) and the National Institute on Drug Abuse (NIDA).

### Author Declarations

The Institutional Review Boards at all COGA sites approved this study. Written consent (and assent for adolescents) was obtained from all participants.

### Summary of Updates

We have added updated analyses, writing, and figures.

## References

1. Agrawal, A., Brislin, S. J., Bucholz, K. K., Dick, D., Hart, R. P., Johnson, E. C., Meyers, J., Salvatore, J., Slesinger, P., Liu, Y., Plawecki, M. H., Kamarajan, C., Pandey, A., Bierut, L., Rice, J., Schuckit, M., Scott, D., Bauer, L., Wetherill, L., … Porjesz, B. (2023). The Collaborative Study on the Genetics of Alcoholism: Overview. *Genes*, Brain and Behavior, 22(5), e12864. 10.1111/gbb.12864

2. Akimova, E. T., Breen, R., Brazel, D. M., & Mills, M. C. (2021). Gene-environment dependencies lead to collider bias in models with polygenic scores. Scientific Reports, 11(1), 1–9. 10.1038/s41598-021-89020-x

3. Baca-Garcia, E., Perez-Rodriguez, M. M., Keyes, K. M., Oquendo, M. A., Hasin, D. S., Grant, B. F., & Blanco, C. (2010). Suicidal ideation and suicide attempts in the United States: 1991-1992 and 2001-2002. Molecular Psychiatry, 15(3), 250–259. 10.1038/mp.2008.98

4. Baurley, J. W., Edlund, C. K., Pardamean, C. I., Conti, D. V., & Bergen, A. W. (2016). Smokescreen: a targeted genotyping array for addiction research. BMC Genomics, 17(1), 145. 10.1186/s12864-016-2495-7

5. Begleiter, H. (1995). The Collaborative Study on the Genetics of Alcoholism. Alcohol Health and Research World, 19(3), 228–236.

6. Bigdeli, T. B., Fanous, A. H., Li, Y., Rajeevan, N., Sayward, F., Genovese, G., Gupta, R., Radhakrishnan, K., Malhotra, A. K., Sun, N., Lu, Q., Hu, Y., Li, B., Chen, Q., Mane, S., Miller, P., Cheung, K.-H., Gur, R. E., Greenwood, T. A., … Harvey, P. D. (2020). Genome-Wide Association Studies of Schizophrenia and Bipolar Disorder in a Diverse Cohort of US Veterans. Schizophrenia Bulletin. 10.1093/schbul/sbaa133

7. Bucholz, K. K., Cadoret, R., Cloninger, C. R., Dinwiddie, S. H., Hesselbrock, V. M., Nurnberger, J. I., Reich, T., Schmidt, I., & Schuckit, M. A. (1994). A new, semi-structured psychiatric interview for use in genetic linkage studies: a report on the reliability of the SSAGA. Journal of Studies on Alcohol, 55(2), 149–158.

8. Cabé, N., Laniepce, A., Ritz, L., Lannuzel, C., Boudehent, C., Vabret, F., Eustache, F., Beaunieux, H., & Pitel, A. L. (2016). Cognitive impairments in alcohol dependence: From screening to treatment improvements. Encephale, 42(1), 74–81. 10.1016/j.encep.2015.12.012

9. Cardenas, V. A., Price, M., & Fein, G. (2018). EEG coherence related to fMRI resting state synchrony in long-term abstinent alcoholics. NeuroImage: Clinical, 17, 481–490. 10.1016/j.nicl.2017.11.008

10. Case, A., & Deaton, A. (2015). Rising morbidity and mortality in midlife among white non-Hispanic Americans in the 21st century. Proceedings of the National Academy of Sciences of the United States of America, 112(49), 15078–15083. 10.1073/pnas.1518393112

11. Colbert, S. M. C., Hatoum, A. S., Shabalin, A., Li, Q. S., Coon, H., Nelson, E. C., Agrawal, A., Docherty, A. R., & Johnson, E. C. (2021). Exploring the genetic overlap of suicide-related behaviors and substance use disorders. *American Journal of Medical Genetics*, Part B: Neuropsychiatric Genetics, 186(8), 445–455. 10.1002/ajmg.b.32880

12. Colin Cameron, A., Gelbach, J. B., & Miller, D. L. (2011). Robust inference with multiway clustering. Journal of Business and Economic Statistics, 29(2), 238–249. 10.1198/jbes.2010.07136

13. Das, S., Forer, L., Schönherr, S., Sidore, C., Locke, A. E., Kwong, A., Vrieze, S. I., Chew, E. Y., Levy, S., McGue, M., Schlessinger, D., Stambolian, D., Loh, P.-R., Iacono, W. G., Swaroop, A., Scott, L. J., Cucca, F., Kronenberg, F., Boehnke, M., … Fuchsberger, C. (2016). Next-generation genotype imputation service and methods. Nature Genetics, 48(10), 1284–1287. 10.1038/ng.3656

14. Davenport, C., Soule, S. A., & Armstrong, D. A. (2011). Protesting while black? the differential policing of american activism, 1960 to 1990. American Sociological Review, 76(1), 152–178. 10.1177/0003122410395370

15. Delaneau, O., Howie, B., Cox, A. J., Zagury, J.-F., & Marchini, J. (2013). Haplotype estimation using sequencing reads. American Journal of Human Genetics, 93(4), 687–696. 10.1016/j.ajhg.2013.09.002

16. Dick, D. M., Balcke, E., McCutcheon, V., Francis, M., Kuo, S., Salvatore, J., Meyers, J., Bierut, L. J., Schuckit, M., Hesselbrock, V., Edenberg, H. J., Porjesz, B., Porjesz, B., Hesselbrock, V., Foroud, T., Agrawal, A., Dick, D., Hesselbrock, V., Edenberg, H. J., … Bucholz, K. (2023). The collaborative study on the genetics of alcoholism: Sample and clinical data. *Genes*, Brain and Behavior, 22(5), e12860. 10.1111/gbb.12860

17. Ding, Y., Hou, K., Xu, Z., Pimplaskar, A., Petter, E., Boulier, K., Privé, F., Vilhjálmsson, B. J., Olde Loohuis, L. M., & Pasaniuc, B. (2023). Polygenic scoring accuracy varies across the genetic ancestry continuum. Nature, 618(7966), 774–781. 10.1038/s41586-023-06079-4

18. Docherty, A. R., Mullins, N., Ashley-Koch, A. E., Qin, X., Coleman, J. R. I., Shabalin, A., Kang, J. E., Murnyak, B., Wendt, F., Adams, M., Campos, A. I., DiBlasi, E., Fullerton, J. M., Kranzler, H. R., Bakian, A. V., Monson, E. T., Rentería, M. E., Walss-Bass, C., Andreassen, O. A., … Ruderfer, D. M. (2023). GWAS Meta-Analysis of Suicide Attempt: Identification of 12 Genome-Wide Significant Loci and Implication of Genetic Risks for Specific Health Factors. American Journal of Psychiatry, 180(10), 723–738. 10.1176/appi.ajp.21121266

19. Dudbridge, F. (2013). Power and Predictive Accuracy of Polygenic Risk Scores. PLoS Genetics, 9(3), e1003348. 10.1371/journal.pgen.1003348

20. Edwards, A. C., Ohlsson, H., Sundquist, J., Crump, C., Mościcki, E., Sundquist, K., & Kendler, K. S. (2024). The role of substance use disorders in the transition from suicide attempt to suicide death: a record linkage study of a Swedish cohort. Psychological Medicine, 54(1), 90–97. 10.1017/S0033291722002240

21. Ehlers, C. L., Wills, D. N., Karriker-Jaffe, K. J., Gilder, D. A., Phillips, E., & Bernert, R. A. (2020). Delta Event-Related Oscillations Are Related to a History of Extreme Binge Drinking in Adolescence and Lifetime Suicide Risk. Behavioral Sciences, 10(10), 154. 10.3390/bs10100154

22. Grant, B. F., Saha, T. D., June Ruan, W., Goldstein, R. B., Patricia Chou, S., Jung, J., Zhang, H., Smith, S. M., Pickering, R. P., Huang, B., & Hasin, D. S. (2016). Epidemiology of DSM-5 drug use disorder results from the national epidemiologic survey on alcohol and related conditions-III. JAMA Psychiatry, 73(1), 39–47. 10.1001/jamapsychiatry.2015.2132

23. Heatherton, T. F., Kozlowski, L. T., Frecker, R. C., & Fagerstrom, K. O. (1991). The Fagerstrom Test for Nicotine Dependence: a revision of the Fagerstrom Tolerance Questionnaire. Br J Addict, 86(9), 1119–1127.

24. Isaacs, J. Y., Smith, M. M., Sherry, S. B., Seno, M., Moore, M. L., & Stewart, S. H. (2022). Alcohol use and death by suicide: A meta-analysis of 33 studies. Suicide and Life-Threatening Behavior, 52(4), 600–614. 10.1111/sltb.12846

25. Johnson, E. C., Salvatore, J. E., Lai, D., Merikangas, A. K., Nurnberger, J. I., Tischfield, J. A., Xuei, X., Kamarajan, C., Wetherill, L., Rice, J. P., Kramer, J. R., Kuperman, S., Foroud, T., Slesinger, P. A., Goate, A. M., Porjesz, B., Dick, D. M., Edenberg, H. J., & Agrawal, A. (2023). The collaborative study on the genetics of alcoholism: Genetics. *Genes*, Brain and Behavior, 22(5), e12856. 10.1111/gbb.12856

26. Kamarajan, C., Ardekani, B. A., Pandey, A. K., Chorlian, D. B., Kinreich, S., Pandey, G., Meyers, J. L., Zhang, J., Kuang, W., Stimus, A. T., & Porjesz, B. (2020). Random forest classification of alcohol use disorder using EEG source functional connectivity, neuropsychological functioning, and impulsivity measures. Behavioral Sciences, 10(3). 10.3390/bs10030062

27. Karlsson Linnér, R., Mallard, T. T., Barr, P. B., Sanchez-Roige, S., Madole, J. W., Driver, M. N., Poore, H. E., de Vlaming, R., Grotzinger, A. D., Tielbeek, J. J., Johnson, E. C., Liu, M., Rosenthal, S. B., Ideker, T., Zhou, H., Kember, R. L., Pasman, J. A., Verweij, K. J. H., Liu, D. J., … Dick, D. M. (2021). Multivariate analysis of 1.5 million people identifies genetic associations with traits related to self-regulation and addiction. Nature Neuroscience, 1–10. 10.1038/s41593-021-00908-3

28. Keilp, J. G., Gorlyn, M., Russell, M., Oquendo, M. A., Burke, A. K., Harkavy-Friedman, J., & Mann, J. J. (2013). Neuropsychological function and suicidal behavior: Attention control, memory and executive dysfunction in suicide attempt. Psychological Medicine, 43(3), 539–551. 10.1017/S0033291712001419

29. Kessler, R. C., Borges, G., & Walters, E. E. (1999). Prevalence of and risk factors for lifetime suicide attempts in the National Comorbidity Survey. Archives of General Psychiatry, 56(7), 617–626. 10.1001/archpsyc.56.7.617

30. Koller, G., Preuß, U. W., Bottlender, M., Wenzel, K., & Soyka, M. (2002). Impulsivity and aggression as predictors of suicide attempts in alcoholics. European Archives of Psychiatry and Clinical Neuroscience, 252(4), 155–160. 10.1007/s00406-002-0362-9

31. Kranzler, H. R., Zhou, H., Kember, R. L., Vickers Smith, R., Justice, A. C., Damrauer, S., Tsao, P. S., Klarin, D., Baras, A., Reid, J., Overton, J., Rader, D. J., Cheng, Z., Tate, J. P., Becker, W. C., Concato, J., Xu, K., Polimanti, R., Zhao, H., … Others. (2019). Genome-wide association study of alcohol consumption and use disorder in 274,424 individuals from multiple populations. Nature Communications, 10(1), 1499. 10.1038/s41467-019-09480-8

32. Kurki, M. I., Karjalainen, J., Palta, P., Sipilä, T. P., Kristiansson, K., Donner, K. M., Reeve, M. P., Laivuori, H., Aavikko, M., Kaunisto, M. A., Loukola, A., Lahtela, E., Mattsson, H., Laiho, P., Della Briotta Parolo, P., Lehisto, A. A., Kanai, M., Mars, N., Rämö, J., … Palotie, A. (2023). FinnGen provides genetic insights from a well-phenotyped isolated population. Nature, 613(7944), 508–518. 10.1038/s41586-022-05473-8

33. Lai, D., Kapoor, M., Wetherill, L., Schwandt, M., Ramchandani, V. A., Goldman, D., Chao, M., Almasy, L., Bucholz, K., Hart, R. P., Kamarajan, C., Meyers, J. L., Nurnberger, J. I., Tischfield, J., Edenberg, H. J., Schuckit, M., Goate, A., Scott, D. M., Porjesz, B., … Foroud, T. (2021). Genome-wide admixture mapping of DSM-IV alcohol dependence, criterion count, and the self-rating of the effects of ethanol in African American populations. *American Journal of Medical Genetics*, Part B: Neuropsychiatric Genetics, 186(3), 151–161. 10.1002/ajmg.b.32805

34. Lannoy, S., Ohlsson, H., Kendler, K. S., Stephenson, M., Sundquist, J., Sundquist, K., & Edwards, A. C. (2024). Risk of suicidal behavior as a function of alcohol use disorder typologies: A Swedish population-based study. Addiction, 119(2), 281–290. 10.1111/add.16351

35. Lannoy, S., Ohlsson, H., Sundquist, J., Sundquist, K., & Edwards, A. C. (2022). Roles of alcohol use disorder and resilience in risk of suicide attempt in men: A Swedish population-based cohort. Suicide and Life-Threatening Behavior, 52(2), 341–351. 10.1111/sltb.12825

36. Le Berre, A. P. (2019). Emotional processing and social cognition in alcohol use disorder. Neuropsychology, 33(6), 808–821. 10.1037/neu0000572

37. Le Berre, A. P., Fama, R., & Sullivan, E. V. (2017). Executive Functions, Memory, and Social Cognitive Deficits and Recovery in Chronic Alcoholism: A Critical Review to Inform Future Research. In Alcoholism: Clinical and Experimental Research (Vol. 41, Issue 8, pp. 1432–1443). Blackwell Publishing Ltd. 10.1111/acer.13431

38. Leuchter, A. F., Cook, I. A., Hunter, A. M., Cai, C., & Horvath, S. (2012a). Resting-state quantitative electroencephalography reveals increased neurophysiologic connectivity in depression. PLoS ONE, 7(2). 10.1371/journal.pone.0032508

39. Leuchter, A. F., Cook, I. A., Hunter, A. M., Cai, C., & Horvath, S. (2012b). Resting-state quantitative electroencephalography reveals increased neurophysiologic connectivity in depression. PLoS ONE, 7(2), e32508. 10.1371/journal.pone.0032508

40. Levey, D. F., Stein, M. B., Wendt, F. R., Pathak, G. A., Zhou, H., Aslan, M., Quaden, R., Harrington, K. M., Nuñez, Y. Z., Overstreet, C., Radhakrishnan, K., Sanacora, G., McIntosh, A. M., Shi, J., Shringarpure, S. S., Concato, J., Polimanti, R., & Gelernter, J. (2021). Bi-ancestral depression GWAS in the Million Veteran Program and meta-analysis in >1.2 million individuals highlight new therapeutic directions. Nature Neuroscience. 10.1038/s41593-021-00860-2

41. Meyers, J. L., Brislin, S. J., Kamarajan, C., Plawecki, M. H., Chorlian, D., Anohkin, A., Kuperman, S., Merikangas, A., Pandey, G., Kinreich, S., Pandey, A., Edenberg, H. J., Bucholz, K. K., Almasy, L., & Porjesz, B. (2023). The collaborative study on the genetics of alcoholism: Brain function. *Genes*, Brain and Behavior, 22(5), e12862. 10.1111/gbb.12862

42. Mills, M. C., & Rahal, C. (2019). A scientometric review of genome-wide association studies. Communications Biology, 2(1), 9. 10.1038/s42003-018-0261-x

43. Modesto-Lowe, V., Brooks, D., & Ghani, M. (2006). Alcohol dependence and suicidal behavior: From research to clinical challenges. In Harvard Review of Psychiatry (Vol. 14, Issue 5, pp. 241–248). 10.1080/10673220600975089

44. Mullins, N., Bigdeli, T. B., Børglum, A. D., Coleman, J. R. I., Demontis, D., Mehta, D., Power, R. A., Ripke, S., Stahl, E. A., Starnawska, A., Anjorin, A., Corvin, A., Sanders, A. R., Forstner, A. J., Reif, A., Koller, A. C., Tkowska, B. S., Baune, B. T., Müller-Myhsok, B., … Lewis, C. M. (2019). GWAS of suicide attempt in psychiatric disorders and association with major depression polygenic risk scores. American Journal of Psychiatry, 176(8), 651–660. 10.1176/appi.ajp.2019.18080957

45. Mullins, N., Forstner, A. J., O’Connell, K. S., Coombes, B., Coleman, J. R. I., Qiao, Z., Als, T. D., Bigdeli, T. B., Børte, S., Bryois, J., Charney, A. W., Drange, O. K., Gandal, M. J., Hagenaars, S. P., Ikeda, M., Kamitaki, N., Kim, M., Krebs, K., Panagiotaropoulou, G., … Andreassen, O. A. (2021). Genome-wide association study of more than 40,000 bipolar disorder cases provides new insights into the underlying biology. Nature Genetics. 10.1038/s41588-021-00857-4

46. Mullins, N., Kang, J. E., Campos, A. I., Coleman, J. R. I., Edwards, A. C., Galfalvy, H., Levey, D. F., Lori, A., Shabalin, A., Starnawska, A., Su, M. H., Watson, H. J., Adams, M., Awasthi, S., Gandal, M., Hafferty, J. D., Hishimoto, A., Kim, M., Okazaki, S., … Willour, V. (2022). Dissecting the Shared Genetic Architecture of Suicide Attempt, Psychiatric Disorders, and Known Risk Factors. Biological Psychiatry, 91(3), 313–327. 10.1016/j.biopsych.2021.05.029

47. Mumtaz, W., Vuong, P. L., Xia, L., Malik, A. S., & Rashid, R. B. A. (2017). An EEG-based machine learning method to screen alcohol use disorder. Cognitive Neurodynamics, 11(2), 161–171. 10.1007/s11571-016-9416-y

48. National Academies of Sciences and Medicine, E. (2023). Using population descriptors in genetics and genomics research: A new framework for an evolving field. In Using Population Descriptors in Genetics and Genomics Research: A New Framework for an Evolving Field. The National Academies Press. 10.17226/26902

49. Nievergelt, C. M., Maihofer, A. X., Klengel, T., Atkinson, E. G., Chen, C. Y., Choi, K. W., Coleman, J. R. I., Dalvie, S., Duncan, L. E., Gelernter, J., Levey, D. F., Logue, M. W., Polimanti, R., Provost, A. C., Ratanatharathorn, A., Stein, M. B., Torres, K., Aiello, A. E., Almli, L. M., … Koenen, K. C. (2019). International meta-analysis of PTSD genome-wide association studies identifies sex- and ancestry-specific genetic risk loci. Nature Communications. 10.1038/s41467-019-12576-w

50. Nunez, P. L., Srinivasan, R., Westdorp, A. F., Wijesinghe, R. S., Tucker, D. M., Silberstein, R. B., & Cadusch, P. J. (1997). EEG coherency. I: Statistics, reference electrode, volume conduction, Laplacians, cortical imaging, and interpretation at multiple scales. Electroencephalography and Clinical Neurophysiology, 103(5), 499–515. 10.1016/s0013-4694(97)00066-7

51. Olfson, M., Blanco, C., Wall, M., Liu, S. M., Saha, T. D., Pickering, R. P., & Grant, B. F. (2017). National Trends in Suicide Attempts Among Adults in the United States. JAMA Psychiatry, 74(11), 1095–1103. 10.1001/jamapsychiatry.2017.2582

52. Pandey, G., Seay, M. J., Meyers, J. L., Chorlian, D. B., Pandey, A. K., Kamarajan, C., Ehrenberg, M., Pitti, D., Kinreich, S., Subbie-Saenz de Viteri, S., Acion, L., Anokhin, A., Bauer, L., Chan, G., Edenberg, H., Hesselbrock, V., Kuperman, S., McCutcheon, V. V., Bucholz, K. K., … Porjesz, B. (2020). Density and Dichotomous Family History Measures of Alcohol Use Disorder as Predictors of Behavioral and Neural Phenotypes: A Comparative Study Across Gender and Race/Ethnicity. Alcoholism: Clinical and Experimental Research, 44(3), 697–710. 10.1111/acer.14280

53. Park, S. M., Lee, J. Y., Kim, Y. J., Lee, J.-Y., Jung, H. Y., Sohn, B. K., Kim, D. J., & Choi, J.-S. (2017). Neural connectivity in Internet gaming disorder and alcohol use disorder: A resting-state EEG coherence study. Scientific Reports, 7(1), 1333. 10.1038/s41598-017-01419-7

54. Peng, Q., Gilder, D. A., Bernert, R. A., Karriker-Jaffe, K. J., & Ehlers, C. L. (2024). Genetic factors associated with suicidal behaviors and alcohol use disorders in an American Indian population. Molecular Psychiatry. 10.1038/s41380-023-02379-3

55. Potash, J. B., Scott Kane, M. H., Chiu, Y., Simpson, S. G., Dean MacKinnon, M. F., McInnis, M. G., McMahon, F. J., & Raymond DePaulo, J. (2000). Attempted Suicide and Alcoholism in Bipolar Disorder: Clinical and Familial Relationships. Am J Psychiatry, 157, 12.

56. Rattana Wannisa, & Rattana Narunart. (2558). H h’.lVJ.16262. Journal, 20(Issue), 5. DOI

57. Richard-Devantoy, S., Berlim, M. T., & Jollant, F. (2014). A meta-analysis of neuropsychological markers of vulnerability to suicidal behavior in mood disorders. Psychological Medicine, 44(8), 1663–1673. 10.1017/S0033291713002304

58. Richard-Devantoy, S., Gorwood, P., Annweiler, C., Olié, J. P., Le Gall, D., & Beauchet, O. (2012). Suicidal behaviours in affective disorders: A deficit of cognitive inhibition? In Canadian Journal of Psychiatry (Vol. 57, Issue 4, pp. 254–262). Canadian Psychiatric Association. 10.1177/070674371205700409

59. Ripley, B., & Venables, W. (2022). Feed-Forward Neural Networks and Multinomial Log-Linear Models. R package Version 7.3-17. https://cran.r-project.org/package=nnet

60. Ruan, Y., Lin, Y. F., Feng, Y. C. A., Chen, C. Y., Lam, M., Guo, Z., Ahn, Y. M., Akiyama, K., Arai, M., Baek, J. H., Chen, W. J., Chung, Y. C., Feng, G., Fujii, K., Glatt, S. J., Ha, K., Hattori, K., Higuchi, T., Hishimoto, A., … Ge, T. (2022). Improving polygenic prediction in ancestrally diverse populations. Nature Genetics, 54(5), 573–580. 10.1038/s41588-022-01054-7

61. Sanchez-Roige, S., Palmer, A. A., Fontanillas, P., Elson, S. L., Adams, M. J., Howard, D. M., Edenberg, H. J., Davies, G., Crist, R. C., Deary, I. J., McIntosh, A. M., Clarke, T.-K., & Clarke, T.-K. (2019). Genome-Wide Association Study Meta-Analysis of the Alcohol Use Disorders Identification Test (AUDIT) in Two Population-Based Cohorts. American Journal of Psychiatry, 176(2), 107–118. 10.1176/appi.ajp.2018.18040369

62. Saunders, G. R. B., Wang, X., Chen, F., Jang, S. K., Liu, M., Wang, C., Gao, S., Jiang, Y., Khunsriraksakul, C., Otto, J. M., Addison, C., Akiyama, M., Albert, C. M., Aliev, F., Alonso, A., Arnett, D. K., Ashley-Koch, A. E., Ashrani, A. A., Barnes, K. C., … Vrieze, S. (2022). Genetic diversity fuels gene discovery for tobacco and alcohol use. Nature 2022 612:7941, 612(7941), 720–724. 10.1038/s41586-022-05477-4

63. Scheer, V., Blanco, C., Olfson, M., Lemogne, C., Airagnes, G., Peyre, H., Limosin, F., & Hoertel, N. (2020). A comprehensive model of predictors of suicide attempt in individuals with panic disorder: Results from a national 3-year prospective study. General Hospital Psychiatry, 67, 127–135. 10.1016/j.genhosppsych.2020.09.006

64. Shepard, D. S., Gurewich, D., Lwin, A. K., Reed, G. A., & Silverman, M. M. (2016). Suicide and Suicidal Attempts in the United States: Costs and Policy Implications. Suicide & Life-Threatening Behavior, 46(3), 352–362. 10.1111/sltb.12225

65. Sher, L. (2006). Alcoholism and suicidal behavior: A clinical overview. In Acta Psychiatrica Scandinavica (Vol. 113, Issue 1, pp. 13–22). 10.1111/j.1600-0447.2005.00643.x

66. Stephenson, M., Lannoy, S., & Edwards, A. C. (2023). Shared genetic liability for alcohol consumption, alcohol problems, and suicide attempt: Evaluating the role of impulsivity. Translational Psychiatry, 13(1), 87. 10.1038/s41398-023-02389-3

67. Tilstra, A. M., Simon, D. H., & Masters, R. K. (2021). Trends in ‘Deaths of Despair’ among Working-Aged White and Black Americans, 1990-2017. American Journal of Epidemiology, 190(9), 1751–1759. 10.1093/aje/kwab088

68. Trubetskoy, V., Pardiñas, A. F., Qi, T., Panagiotaropoulou, G., Awasthi, S., Bigdeli, T. B., Bryois, J., Chen, C. Y., Dennison, C. A., Hall, L. S., Lam, M., Watanabe, K., Frei, O., Ge, T., Harwood, J. C., Koopmans, F., Magnusson, S., Richards, A. L., Sidorenko, J., … van Os, J. (2022). Mapping genomic loci implicates genes and synaptic biology in schizophrenia. Nature, 604(7906), 502–508. 10.1038/s41586-022-04434-5

69. Walters, R. K., Polimanti, R., Johnson, E. O. E. C. E. O., McClintick, J. N., Adams, M. J., Adkins, A. E., Aliev, F., Bacanu, S.-A., Batzler, A., Bertelsen, S., Biernacka, J. M., Bigdeli, T. B., Chen, L.-S., Clarke, T.-K., Chou, Y.-L., Degenhardt, F., Docherty, A. R., Edwards, A. C., Fontanillas, P., … Agrawal, A. (2018). Trans-ancestral GWAS of alcohol dependence reveals common genetic underpinnings with psychiatric disorders. Nature Neuroscience, 21(12), 1656–1669. 10.1038/s41593-018-0275-1

70. Wang, J.-C., Foroud, T., Hinrichs, A. L., Le, N. X. H., Bertelsen, S., Budde, J. P., Harari, O., Koller, D. L., Wetherill, L., Agrawal, A., Almasy, L., Brooks, A. I., Bucholz, K., Dick, D., Hesselbrock, V., Johnson, E. O., Kang, S., Kapoor, M., Kramer, J., … Goate, A. M. (2013). A genome-wide association study of alcohol-dependence symptom counts in extended pedigrees identifies C15orf53. Molecular Psychiatry, 18(11), 1218–1224. 10.1038/mp.2012.143

71. Whiteford, H. A., Degenhardt, L., Rehm, J., Baxter, A. J., Ferrari, A. J., Erskine, H. E., Charlson, F. J., Norman, R. E., Flaxman, A. D., Johns, N., Burstein, R., Murray, C. J., & Vos, T. (2013). Global burden of disease attributable to mental and substance use disorders: findings from the Global Burden of Disease Study 2010. The Lancet, 382(9904), 1575– 1586. 10.1016/S0140-6736(13)61611-6

72. Yuodelis-Flores, C., & Ries, R. K. (2015). Addiction and suicide: A review. In American Journal on Addictions (Vol. 24, Issue 2, pp. 98–104). Wiley-Blackwell Publishing Ltd. 10.1111/ajad.12185

73. Zatti, C., Rosa, V., Barros, A., Valdivia, L., Calegaro, V. C., Freitas, L. H., Ceresér, K. M. M., Rocha, N. S. da, Bastos, A. G., & Schuch, F. B. (2017). Childhood trauma and suicide attempt: A meta-analysis of longitudinal studies from the last decade. Psychiatry Research, 256, 353–358. 10.1016/j.psychres.2017.06.082

74. Zhou, H., Kember, R. L., Deak, J. D., Xu, H., Toikumo, S., Yuan, K., Lind, P. A., Farajzadeh, L., Wang, L., Hatoum, A. S., Johnson, J., Lee, H., Mallard, T. T., Xu, J., Johnston, K. J. A., Johnson, E. C., Nielsen, T. T., Galimberti, M., Dao, C., … Gelernter, J. (2023). Multi-ancestry study of the genetics of problematic alcohol use in over 1 million individuals. Nature Medicine, 29(12), 3184–3192. 10.1038/s41591-023-02653-5

75. Zhou, H., Sealock, J. M., Sanchez-Roige, S., Clarke, T. K., Levey, D. F., Cheng, Z., Li, B., Polimanti, R., Kember, R. L., Smith, R. V., Thygesen, J. H., Morgan, M. Y., Atkinson, S. R., Thursz, M. R., Nyegaard, M., Mattheisen, M., Børglum, A. D., Johnson, E. C., Justice, A. C., … Gelernter, J. (2020). Genome-wide meta-analysis of problematic alcohol use in 435,563 individuals yields insights into biology and relationships with other traits. Nature Neuroscience. 10.1038/s41593-020-0643-5

