## supplemental info for "Clinical, genomic, and neurophysiological correlates of lifetime suicide attempts among individuals with an alcohol use disorder"

### Genome Wide Association Study (GWAS) of Suicide Attempt in Participants with an Alcohol Use Disorder

We conducted GWAS, on 7,784,968 SNPs in the EUR sample and 16,100,604 SNPs in the AFR sample using a mixed model incorporating a genetic relationship matrix to control for relatedness [1] in the *GWAF* package in R [2]. We included sex, age, the first three genetic principal components (PCs), genotype array, and birth cohort (prior to 1930, 1930-1949, 1950-1969, and 1970 and after) as covariates. GWAS were stratified by genetic similarity, using identical phenotypic definitions, covariates, SNP QC standards, MAF thresholds and imputation protocols. Subsequently, we meta-analyzed across the AFR and EUR results using inverse-variance fixed-effects weighting and genomic control in METAL [3]. We used established thresholds for genome wide significance (p< 5 x 10^-8^). We also conducted a post-hoc GWAS analysis covarying for depression, given the high prevalence.

Next we performed a series of post-GWAS analyses using a protocol outlined in previous analyses [4]. We limited results to the EUR only given the small sample size of the AFR analyses and the lack of AFR predicted transcriptomic expression results in some of the post-GWAS pipelines. To identify functional enrichment, we used MAGMA software (version 1.08), and its recent intersessions (FUMA version 1.3.6) [5], a method for gene-level and gene-set enrichment analysis using GWAS summary statistics. In all the MAGMA-based analyses, SNPs were annotated to the 20,260 coding genes from Ensembl v92, with a 1 kb window for both sides (i.e., start and end). Since GWAS contained EUR and AFR samples, we used the 1000G European and African panels [6] respectively to account for linkage disequilibrium (LD) between SNPs. Finally, we corrected all tests for multiple-testing using a Bonferroni correction.

Next, we used the summary-data-based Mendelian randomization (SMR) method to test for a joint association between GWAS summary statistics SNPs and eQTL, using the default settings in the SMR software [7] and the 1000G European ancestries reference panel. We again applied a Bonferroni correction for multiple-testing on the SMR P-value (PSMR). Moreover, a post-filtering step was applied by conducting heterogeneity in dependent instruments (HEIDI) test. The HEIDI test distinguishes the causality and pleiotropy models from the linkage model by considering the pattern of associations using all the SNPs that are significantly associated with gene expression in the cis-eQTL region. The null hypothesis is that a single variant is associated with both trait and gene expression, while the alternative hypothesis is that trait and gene expression are associated with two distinct variants. We defined significant hits based on SMR-HEIDΙ as those for which PSMR met the Bonferroni significance threshold and had PHEIDI>0.05.

Lastly, we used the JEPEGMIX2-P software [8] with default settings to conduct TWAS using only the 13 brain-specific GReX models coming from GTeX v8 [9]. This method was preferable since it relied on a covariance matrix based on 33K samples compared to other TWAS methods which use less than 3k samples. We applied the within-tissue Bonferroni correction to detect significant TWAS genes.

Within the analytic sample we performed a GWAS of SA in those with available genetic data (no SA = 2,495 EUR and 643 AFR; SA = 775 EUR and 155 AFR). There was no individual SNP associated with suicide attempts that reached genome-wide significance (see supplemental information for full results). For the post-GWAS analyses, there were no significant gene-based or gene-set enrichment from the MAGMA results (see supplemental tables S5 – S6). Additionally, none of the results from the SMR analyses reached significance after correcting for multiple testing (see supplemental tables S7 – S9).

Supplemental Figure 1: Suicide GWAS Results

SNP based genome-wide association study findings comparing COGA participants with AUD who have attempted suicide (N: 930) compared to those who have not attempted suicide (N: 3,138), in the (top) multi-ancestry meta-analysis, (middle) European-like specific GWAS, and (bottom) African-like specific GWAS.

### Neuropsychological tasks

In an exploratory series of analyses, we compared those with alcohol use disorder (AUD) who attempted suicide and those with AUD who did not attempt suicide across a battery of neuropsychological measures on a subset of COGA participants (*N* = 188). Neuropsychological tasks included the Tower of London Task (TOLT) and the Visual Span Task (VST). These tasks have been detailed in previous publications [10]. Briefly, as part of the Colorado assessment tests for cognitive and neuropsychological assessment [11], the TOLT assesses planning and problem-solving ability. Participants are asked to solve a set of puzzles with graded difficulty levels by arranging the color beads one at a time from a starting position to a desired goal position in as few moves as possible. Participant performance on the TOLT was defined in the current study by the number of optimal trials achieved. The VST, also part of the Colorado assessment tests [11], requires subjects to duplicate a pattern of sequentially illuminated stimuli as well as to generate that pattern in reverse order. This test was used to assess visuospatial memory span from the forward condition and working memory from the backward condition. Participant performance on the VST was defined in the current study by the total forward and backward span (maximum sequence-length achieved).

We used multiple-group, multi-level regression models conducted in Mplus [12]. We included sex, age (at time of neuropsychological or neurophysiological assessment as appropriate), genetic similarity, family history of AUD [13], and family relatedness as covariates. Since neuropsychological task performance was evaluated longitudinally, we used the most recent assessment from each individual (mean age = 24.2; SD = 12.3). We observed differences in neuropsychological task performance differences among AUD individuals who had attempted suicide. Those with AUD who reported a lifetime suicide attempt had fewer optimal trials in the Tower of London Task (an average of 18 optimal trials among those without suicide attempts as compared with 17 optimal trials among those with suicide attempt, *p* < 0.05) and both a shorter attention span and short-term memory span in the Visual Span Task (an average of 10 words among those without suicide attempts as compared with 9 words among those with suicide attempt, *p* < 0.05). There were no significant differences between those with and without a lifetime suicide attempt in the Tower of London Task *trial time* or Visual Span Task *backward span*. These results point to some differences in cognition across suicide attempt within those with AUD, but further work is necessary to determine whether these exploratory results will replicate.

### References

1.	Lai D, Wetherill L, Bertelsen S, Carey CE, Kamarajan C, Kapoor M, et al. Genome-wide association studies of alcohol dependence, DSM-IV criterion count and individual criteria. Genes Brain Behav. 2019;18.

2.	Chen MH, Yang Q. GWAF: An R package for genome-wide association analyses with family data. Bioinformatics. 2009;26:580–581.

3.	Willer CJ, Li Y, Abecasis GR. METAL: fast and efficient meta-analysis of genomewide association scans. Bioinformatics. 2010;26:2190–2191.

4.	Chatzinakos C, Pernia CD, Morrison FG, Iatrou A, McCullough KM, Schuler H, et al. Single-Nucleus Transcriptome Profiling of Dorsolateral Prefrontal Cortex: Mechanistic Roles for Neuronal Gene Expression, Including the 17q21.31 Locus, in PTSD Stress Response. Am J Psychiatry. 2023;180:739–754.

5.	Watanabe K, Taskesen E, van Bochoven A, Posthuma D. Functional mapping and annotation of genetic associations with FUMA. Nat Commun. 2017;8:1–11.

6.	Auton A, Abecasis GR, Altshuler DM, Durbin RM, Bentley DR, Chakravarti A, et al. A global reference for human genetic variation. Nature. 2015;526:68–74.

7.	Zhu Z, Zhang F, Hu H, Bakshi A, Robinson MR, Powell JE, et al. Integration of summary data from GWAS and eQTL studies predicts complex trait gene targets. Nat Genet. 2016;48:481–487.

8.	Chatzinakos C, Georgiadis F, Lee D, Cai N, Vladimirov VI, Docherty A, et al. TWAS pathway method greatly enhances the number of leads for uncovering the molecular underpinnings of psychiatric disorders. American Journal of Medical Genetics Part B: Neuropsychiatric Genetics. 2020;183:454–463.

9.	Consortium TGte, Aguet F, Anand S, Ardlie KG, Gabriel S, Getz GA, et al. The GTEx Consortium atlas of genetic regulatory effects across human tissues. Science (1979). 2020;369:1318–1330.

10.	Subbie-Saenz de Viteri S, Pandey A, Pandey G, Kamarajan C, Smith R, Anokhin A, et al. Pathways to post-traumatic stress disorder and alcohol dependence: Trauma, executive functioning, and family history of alcoholism in adolescents and young adults. Brain Behav. 2020;10.

11.	Davis HP KFR. Colorado Springs: Colorado Assessment Tests. 1998. Colorado Assessment Test manual. 1998.

12.	Muthén LK, Muthén B. Mplus. The Comprehensive Modelling Program for Applied Researchers: User’s Guide. 2016. 2016.

13.	Pandey G, Seay MJ, Meyers JL, Chorlian DB, Pandey AK, Kamarajan C, et al. Density and Dichotomous Family History Measures of Alcohol Use Disorder as Predictors of Behavioral and Neural Phenotypes: A Comparative Study Across Gender and Race/Ethnicity. Alcohol Clin Exp Res. 2020;44:697–710.
